# Exploration of using “distance-to-bound” to manipulate the difficulty during motor imagery BCI training after stroke – A clinical two-cases study

**DOI:** 10.1101/2025.11.05.25339460

**Authors:** Jonatan Tidare, Martin Johansson-Alvarez, Jeanette Plantin, Susanne Palmcrantz, Elaine Astrand

## Abstract

**Objective:** Motor Imagery-based Brain-Computer Interfaces (MI-BCIs) is a promising technology for neurorehabilitation after stroke. However, many face challenges in using a BCI because they fail to produce discriminable patterns in their brain activity. Personalizing the BCI task difficulty could help the learning process of these users but there is currently very limited knowledge on which methods can be used online. Our aim was to explore a distance-to-bound approach for adapting MI BCI task difficulty in real time.

**Approach:** Two chronic stroke patients performed 12 BCI training sessions over 4 weeks during which they performed MI of open– and close hand movements and received continual visual feedback based on multivariate decoding of ongoing electroencephalogram (EEG) activity. We increased the difficulty and maintained it by adapting it in real time based on distance-to-bound decoding metrics and using a multiple-session design we investigated the stability of this approach and how it related to MI-related EEG activity of each patient.

**Main results:** We show that patients had to produce stronger alpha and beta event-related desynchronization (ERD) activity across the sensorimotor cortical areas of the brain to receive positive feedback. In addition, we show that the online adaptation converged within sessions as well as accommodating for drift in the data both within and between sessions. We suggest that the distance-to-bound approach can effectively be used to control BCI task difficulty and potentially guide patients to produce functionally relevant activity patterns. However, from our results, stronger sensorimotor ERD activity did not consistently correlate to improved motor function. Clinical assessments showed that both patients improved in motor function (+4 and +8.7 change in Fugl-Meyer assessment for upper extremity), however, the correlation to sensorimotor ERD activity was positive for one patient and negative for the other (Pearson’s rho = 0.95,-0.80, p = 0.05, 0.18). These results indicate that the translation of distance-to-bound outputs to feedback needs to be individually tailored considering the stroke lesion and EEG activity profiles for each patient.

**Significance:** This study provides valuable insights and considerations for BCI difficulty adaptation in the aim of developing more effective training protocols in BCI-based stroke rehabilitation.

**Trial registration:** The study was registered at clinicaltrials.gov (NCT03994042) and complied with local rules and regulations according to the Swedish Ethics Review Authority (dnr. 2019-01577).

## Introduction

Motor Imagery (MI), the mental rehearsal of motor tasks, has emerged in stroke rehabilitation as a promising therapeutic activity to promote brain plasticity and improve physical recovery (1). However, MI is difficult to perform and adapt to activity-based therapy due to the lack of visible feedback (2). The brain activity can be recorded using electroencephalography (EEG) during MI, and by using multivariate decoding tools, information related to MI can be extracted in real time (3). Thereby using a brain-computer interface (BCI), a system able to decode mental tasks from ongoing brain activity and provide real-time feedback, can facilitate the performing of MI. As such, recorded brain activity is analysed and interpreted in real-time to produce an output either used to control an external device (such as for example a robotic exoskeleton or orthosis (4)) or provide feedback on brain activity (i.e. neurofeedback). Training with a BCI is a promising technology for neurorehabilitation (5) as it can reinforce learning of controlling an output (i.e. feedback) and consequently modify the underlying brain activity through operant conditioning (6). The theoretical underpinning of MI-based BCI (MI-BCI) rehabilitation is to close the loop between cortical activity (MI-related brain activity patterns) and overt feedback to enable a neural learning environment that can restore, strengthen as well as to create new functional connections in the brain. During the use of a BCI, positive feedback is provided when the performance of MI induces functionally relevant brain activity and, indeed, training MI with feedback after stroke has shown promising outcomes (7,8). Nevertheless, MI-BCIs in stroke research face many challenges, among the most important being the replicability in producing clinically relevant outcomes and the high proportion of non-performers/non-learners (often referred to as ‘BCI illiterates’). Personalization of BCI training might be key in addressing these challenges. Specifically, the extraction of individual brain features common to both MI and motor recovery, and the generation of feedback that is adaptive and aligned to the performance level of the user are examples of personalization strategies that may be critical for learning to use a BCI.

Rather than extracting univariate features of brain activity to represent MI, distributed patterns of activity can be identified using multivariate pattern analysis to establish a statistical relationship between performing MI and simultaneous brain activity (9). By using similar algorithms to classify brain activity patterns, one can also decode ongoing mental processes, such as MI, from the brain activity. Since this approach better accounts for individual variability in brain activity, it is frequently used in BCI research to extract real-time information related to MI performance, which is then transformed into feedback to close the loop (10). Further, common practice when using this decoding approach is to provide feedback based on a discretized output (e.g. MI versus an idle task) from the classification algorithm (11). However, the classification output is often a continuous score or probability value representing certainty of the prediction or distance to a hyperplane (12). The distribution of classifier predictions or scores and how this distribution changes within or between sessions can indicate drift in brain signals and this metric could potentially be used for online adaptation of a classifier to account for drift (13). In the case of linear classifiers, they represent the boundary between classes with a hyperplane in the feature space, and the prediction consists of evaluating on which side of the hyperplane a data sample is located. In this case, the distance to the hyperplane could contain more information than the prediction, known as the distance-to-bound approach (14). If EEG feature amplitudes correlate with strength or decidedness of mental tasks, similar to how it has been shown that imagining different force loads correlate with EEG feature amplitudes (15), then the distance-to-bound approach could model MI behaviour and its relative strength (11). It might therefore open the possibility to adapt the difficulty of the BCI task to facilitate learning and accommodate for individual differences in learning rate. However, little is known of the distance-to-bound approach in BCI rehabilitation, and whether it can used to adapt the difficulty in the objective of promoting stronger functionally relevant MI-patterns.

In this study, we set out to explore the distance-to-bound approach by adapting online, the feedback threshold (FBT) used to determine positive versus negative feedback. The overall difficulty of the BCI task was increased by setting the FBT to generate approximately 40% positive feedback for each patient. This FBT was then also adapted online to account for learning-related effects and maintain the predefined difficulty level. Two stroke patients conducted multiple BCI training sessions with online adaptation of the FBT. From the data that was collected, we applied intra-patient between-session analyses to investigate the stability of the FBT updates within sessions and how the FBT adaptation accommodated for drift in the EEG signals both within and between sessions. Further, we investigated how the FBT was related to cortical activity patterns related to MI.

Our findings show that stronger ERDS patterns for alpha and beta frequency bands across the sensorimotor cortex was required to receive positive feedback for both patients during the BCI training. These activity patterns are consistent with MI-related patterns found in the literature (16,17). We further show that the online adaption of the FBT followed within– and between-session drift in the EEG signals. Importantly, we observe that the functional relevance of producing stronger alpha and beta ERDS across the sensorimotor cortex is not straightforward and plausibly depend on the lesion and the symptomatic EEG activity profile (e.g. under vs. overactivation).

## Materials and Method

Two patients with chronic ischemic stroke in the left hemisphere (stroke occurring >6 months prior to study entry) participated in this study: 1 female and 1 male, aged in their 70’s and 30’s. The extent and location of their lesions are described using Magnetic Resonance Imaging (MRI) and EEG in figure 1. Both patients had lesions within the left middle cerebral artery (MCA) distribution. Lesion volume measured 110cc and 38cc respectively, involving both cortical and subcortical structures. Specifically, patient 1 had a larger lesion spanning gray matter areas including pre– and postcentral gyrus and the paracentral lobule (S1/M1) as well as the superior frontal gyri (premotor cortex). White matter areas overlapping with the lesion of patient 1 included superior and posterior corona radiata and the superior longitudinal fasciculus. Patient 2 had a more focal lesion spanning gray matter areas including the basal ganglia and insula and white matter areas including anterior and superior parts of the corona radiata and the anterior and posterior limb of the internal capsule. Focal slowing (18) of EEG activity can be observed over lesioned areas as increased power in the frequency bands Delta, Theta and Alpha during MI and idle trials of the calibration sessions. Both patients displayed difficulties opening and closing their affected hand and scored 39 and 41 (excluding reflex items) on the motor function score included in the Fugl-Meyer Assessment for Upper Extremity (FMA-UE) prior to study entry (19). The patients’ spasticity was measured using the Neuroflexor method (20) (neural component = 6.12, 17.5), indicating moderate to severe hand spasticity. Cognitive impairment was measured with Montreal Cognitive Assessment (MoCA = 19, 28), indicating the presence of cognitive impairment in patient 1 (21). Neither patient suffered from spatial neglect (Baking tray task = 8, 8) (22). Overall stroke severity was measured using the National Institutes of Health Stroke Scale (NIHSS = 4, 2) indicating a mild stroke (23). Inclusion/exclusion criteria are detailed in the supplementary material.

**Figure 1.**
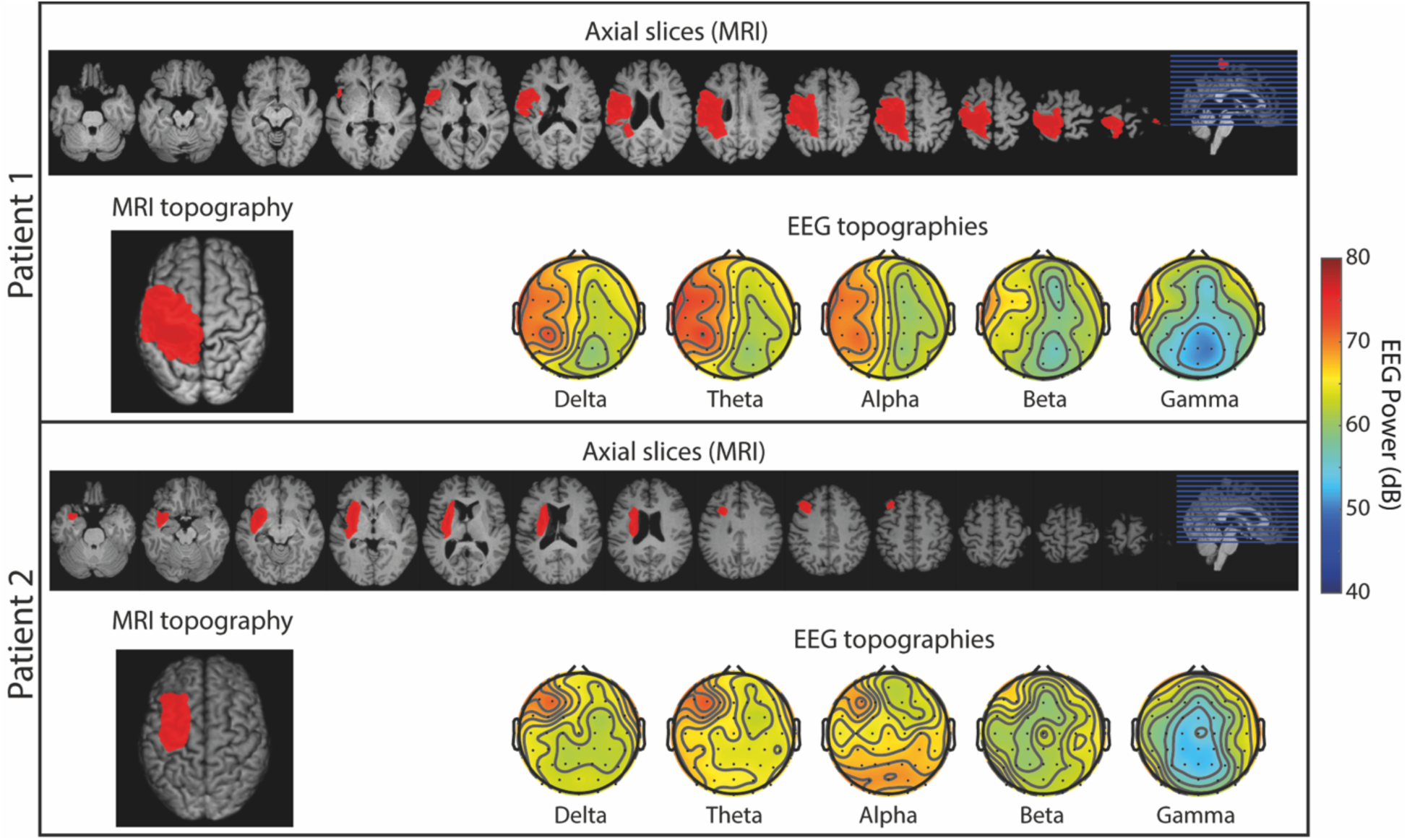
Lesion extent and location for each patient. Manually segmented lesion volumes (red areas in MRI topographies) overlaid on axial slices of template normalized T1-weighted Magnetic Resonance Imaging (MRI) images. Lesion distribution corresponded to middle cerebral artery territory. Electroencephalogram (EEG) topographies show absolute power, averaged across MI and idle trials of the calibration sessions for each patient, in frequency bands Delta, Theta, Alpha, Beta, and Gamma, respectively.

## Study design and user tasks

### Intervention outline

Each patient performed a multiple week intervention consisting of three phases: calibration/baseline, BCI training, and evaluation, led by a trained BCI operator. During calibration and evaluation sessions, EEG was recorded without feedback being provided to the patients. During the BCI training phase, the patient received real-time feedback on the screen while performing MI and idle tasks. Clinical tests were performed throughout the entire intervention, and structural and functional MRI, were measured before and after the BCI training phase. The precise schedule for each patient is described in supplementary figure 1.

### Clinical tests

During each clinical test session, patients were assessed by an experienced physiotherapist and performed a battery of tasks to evaluate hand spasticity, stroke severity, cognitive impairment, spatial neglect, and motor function of arm and hand. In this study, we focus on motor function, measured with FMA-UE motor score (where a max score of 60 points, excluding reflex items, indicates no motor impairment) and the hand subscore (where a max score of 14 points indicate no impairment) (24) as the other assessments are out of the scope for the aim of this study. Clinically important difference for chronic stroke patients with moderate hemiparesis ranges from 4.25 to 7.25 on the FMA-UE (25).

### Neuroimaging

MRI using whole brain spin-echo T1 and T2 weighted sequences was performed to provide high resolution anatomical images for description of lesion size and location. The measurement lasted for approximately 25 minutes and patients were instructed to lay still and relax. All MRI sessions were performed using a Siemens MAGNETOM Prisma 3T scanner. The measurements were made without contrast (non-invasive). Each patient filled out a questionnaire and was screened for metal objects before each MRI session. Lesion maps were manually drawn on all axial slices of native anatomical images (from T1) using MRIcron (26). Lesion location was verified on diffusion images and lesion maps were binarized. The delineation was performed by author J.P., experienced in lesion segmentation, and a neurologist verified lesion maps. For the scope of this study, only structural MRI will be considered.

### EEG calibration

The patients were instructed to perform repeated trials of two tasks: MI of opening or closing the affected hand, and an idle task. Patients were instructed to perform kinaesthetic MI, as opposed to visual MI that has been shown to produce weaker EEG motor activation (27). Each trial started with an animated instruction (visual and auditory) lasting 4-6 seconds, followed by a task onset (hand outline highlighted). After the task onset, the patient began performing the task for a duration of 4-6 seconds. There were 8 seconds of inter-trial pause between each trial, and trial order was pseudo-randomized. A total of 360 trials were recorded per patient, split evenly between 3 calibration sessions (180 MI and 180 idle). During the calibration trials, patients also performed trials of attempted Motor Execution (ME) of the hand task before each MI task to facilitate the imagination of hand movements. The EEG data of the attempted hand movement was excluded from the analysis in this study.

### BCI training

Patients first performed a short calibration block consisting of 32 trials, task identical to the calibration sessions. This block was followed by 80 trials (40 MI, 40 Idle) during which real-time BCI feedback was presented on the computer screen. The trial structure was identical to the calibration sessions except for 1) no ME was performed, and 2) the duration of task performance lasted 10-12 seconds. See figure 2A for an illustration of the BCI trial composition. The BCI feedback was based on the output from a Support Vector Machine (SVM) classifier. The feedback consisted of a moving hand displayed on a computer screen and was updated every 100 ms throughout the feedback trial, starting at 250 ms after task onset until the end of trial. This means that the 1^st^ feedback was based on data from 250 ms–500 ms after task onset, the 2^nd^ feedback was based on data from 350 ms–600 ms after task onset, etc. In each trial, the hand started in a neutral position and the classifier predictions caused the hand to move towards fully open or fully closed depending on the MI task. To prevent a jittering hand, two sequential identical classifier predictions were required to change direction of the moving hand (one prediction was required to continue moving in the same direction). There were 16 steps to reach a fully open or closed hand position (requiring a minimum of 1.6 seconds). Feedback was provided both during MI and idle trials. During idle trials, one direction (open or close) was randomly chosen and patients were instructed to remain in an idle state to maintain the hand in its neutral position.

**Figure 2.**
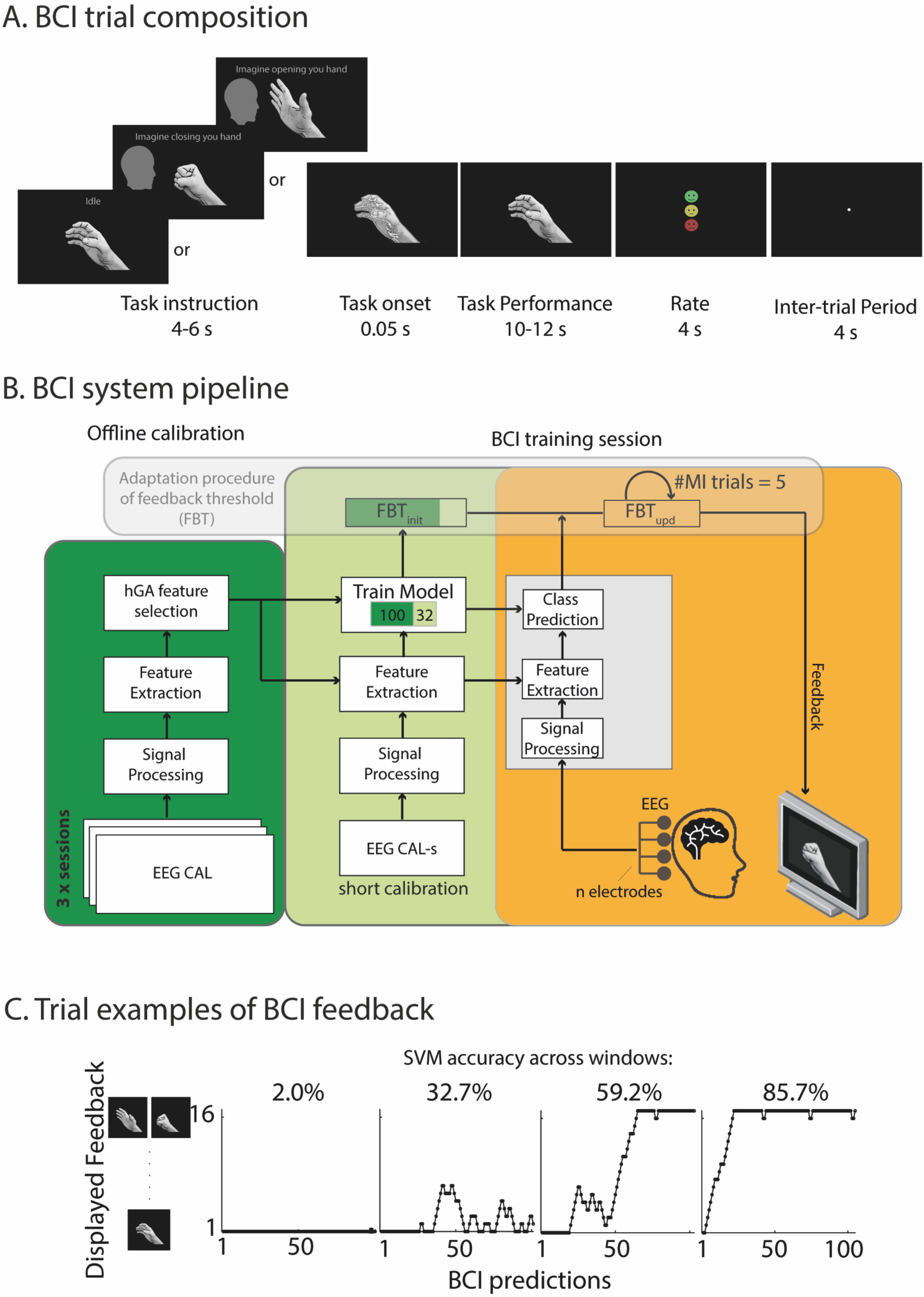
Study design. A) BCI trial composition. Each trial was recorded by first presenting a task instruction (MI open, MI close, or idle), after which the contour around the hand briefly lit up, hence signaling the task onset. This was followed by 10-12 seconds of real-time feedback (i.e. computer hand movements) during MI task performance. Last, a rating and inter-trial period of 4 seconds followed. B) BCI system pipeline. The BCI system was calibrated on data recorded during the three calibration sessions (dark green square) and hGA features used during BCI training were selected based on this data. Each BCI training session began with a short calibration block of 32 trials (light green) of which the data was input together with 100 randomly drawn trials from the calibration sessions, to the retraining of an SVM model. During BCI training (yellow square), the SVM model predicted new data and provided real-time feedback to the patients based on each prediction. The feedback threshold (FBT) was set to reflect the 60^th^ percentile of the SVM score distribution of MI trials and it was updated every 5 MI trials using all available data. C) Trial examples of BCI feedback. The feedback from four MI trials is presented, providing examples of how different levels of single-trial classification accuracy (2.0%, 32.7%, 59.2% and 85.7%) translated into feedback as displayed to patients. The y-axis has 16 steps, ranging from neutral hand to fully opened or fully closed. The x-axis has approximately 110 points representing BCI predictions. A graph reaching the top of the y-axis corresponds to a fully opened or closed hand on the screen.

Prior to each session (including calibration, BCI training and evaluation sessions), we addressed overt artifacts by meticulous instructions and by cautiously monitoring patients and EEG, EMG and accelerometer signals during the intervention. Prior to each session, the test operator thoroughly explained and showed the patients the EEG signal in real time. The operator asked them to do slight movements, yawn, or blink while observing the EEG raw data together, to clarify that these small movements cause highly visible noise in the EEG data. Detailed instructions to relax, sit still and to avoid blinking and body movements during task performance, as well as to look straight forward during both MI and idle tasks and not look at their real hand, was provided orally to patients prior to each session.

The most critical type of noise that could negatively affect this experiment is task-related noise, i.e. non-static noise. Class-related noise can be generated if the patient for example focuses their gaze more strictly during MI, while relaxing their gaze during idle tasks or systematically produces a small movement during MI. We addressed task-related noise during the experiment by active monitoring of patients during the intervention. If systematic movements or artifacts were detected during a session, the task was paused, and the operator communicated with the patients directly. The noisy trial was then re-run as the operator resumed recording. Second, we also carefully examined EMG, accelerometer signals (supplementary figure 2) and the selected EEG features to identify potential artefactual features (figure 3).

**Figure 3.**
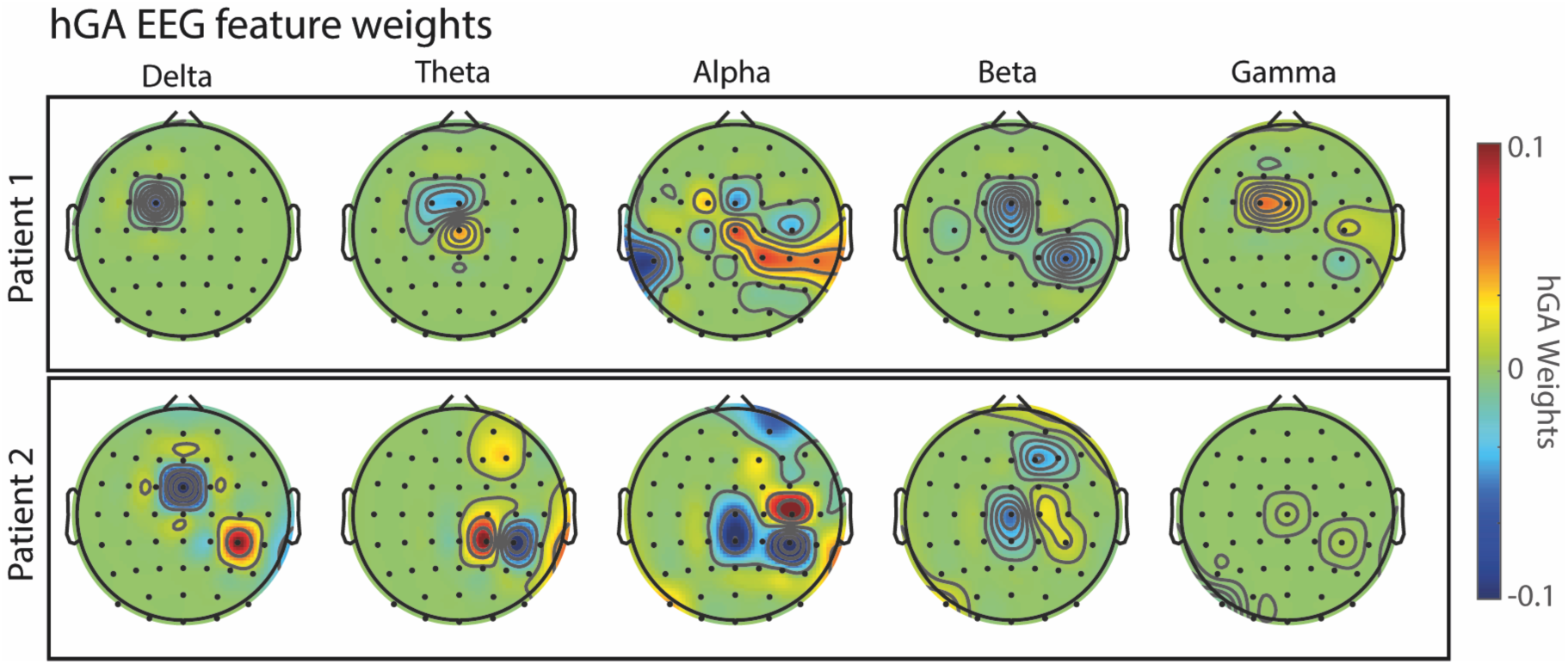
hGA EEG feature weights. Topographies illustrate weights of the selected hGA features for each frequency band (averaged within the frequency band) averaged across the 12 classifiers used during the intervention. Positive (red) weights indicate that MI is represented by increased EEG power relative to idle, while negative (blue) weights indicate that MI is represented by decreased EEG power relative to idle. Zero weight (green) indicate that the feature was not selected by the hGA.

## Data acquisition

EEG, electromyogram (EMG), electrooculogram (EOG), and accelerometer data were synchronously sampled at 1 kHz using a commercial amplifier (actiCHamp, Brain Products). EEG was recorded and sampled using 62 Ag/AgCl active electrodes (Brain Products) positioned according to the international 10/10 standard. Impedances were below 30 kOhm before and after each session, hence below the 40 kOhm limit recommended by the manufacturer. The ground electrode was placed at Fpz and the reference was placed on the left nose wing.

Passive Ag/AgCl electrodes were used to record two channels of EMG and one channel of horizontal EOG. EMG was recorded from the right forearm. Two electrodes were placed 2 cm apart on the medial group of antebrachial muscles with a reference electrode placed on the medial epicondyle, and two were placed on the lateral group of antebrachial muscles with a reference electrode placed on the lateral epicondyle. For horizontal EOG, two electrodes were placed on each temple and the reference on the left earlobe. For vertical EOG, one Ag/AgCl active electrode was placed below the right eye and compared with FP2.

Accelerometer data was recorded using two 1D acceleration sensors (Brain Products), placed on the back of the right hand and the second one on the intermediate phalanx of the middle finger. EMG and accelerometer data during EEG sessions are presented in supplementary figure 2. Additionally, photosensor data (Brain Products) was recorded from a sensor placed on the top-right corner of the patient screen. Every time the computer screen was updated with a task event, the area directly below the photosensor would switch colour (black/white), thus allowing the recording of precise event time stamps.

## EEG signal processing and feature extraction

Raw EEG data were extracted from the task performance part of each trial using overlapping windows of 250 ms window length and 100 ms step size (150 ms overlap). To remove drift in the signal, the DC level was removed by subtracting the average, and surface Laplacian spatial filter was applied to reduce the effect of volume conduction. Power spectral density was calculated at 59 linearly spaced frequencies (2, 3, …, 60 Hz) as the absolute values from the fast Fourier transform (zero-padded to 1000). 62 channels and 59 frequency levels resulted in 3658 features. The time series for each feature was minmax-normalized, using the 90^th^ percentile instead of max to avoid the impact of outliers.

## Feature reduction and classification procedure

The overall process for BCI calibration and training is shown in figure 2B. The number of features was reduced prior to BCI training by using an hierarchical genetic algorithm (hGA, (28–30)) based on Minimum Redundancy Maximum Relevance (mRMR) (31). We modified the hGA in (30) to not consider time windows (i.e. we removed the third hierarchical level). First, a population of 15 different random solutions/individuals were created with two levels of hierarchy; its *n* channels, X = { *x*_1_, *x*_2_, …, *x*_*i*_, …, *x*_*n*_} where *x*_*i*_ ∈ {0, 1} depending on if the channel was selected or not, and its *m* frequencies, Y =. *y*_1_, *y*_2_, …, *y*_j_, …, *y*_*m*_0 where *y*_j_ ∈ {0, 1}. From the initial populations, GA performs 4 steps to create new individuals (offsprings): parent selection, crossover, mutation, and population update. We estimated the fitness by calculating two measures: the correlation between the selected features (CFS) and the percentage of the reduction of the features (PRF). The fitness for the ith individual, *F*_*i*_, was calculated by using equations 1-3:

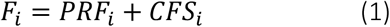

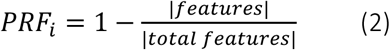

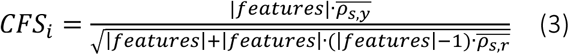

Where |*features*| is the number of features selected, |*total features*| is the total number of features, 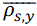 is the average value of the Pearson’s correlation coefficient^1^ between the features and the classes (here MI and idle), and 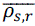 the average value of the Pearson’s correlation coefficient^1^ of every feature to each other. Due to the hierarchy, crossover and mutation was calculated for each level separately. In the crossover step, probabilities were set to 0.5 for both channels and frequencies. In the mutation step, probabilities were set to 0.05 for channels and 0.01 for frequencies. The number of offsprings per generation was set to 4. The offspring always replaced its worst parent if its fitness was higher. The maximum number of evaluations was 40000 (due to the generation limit of 10000), with a break point if fitness value ever reached 2.

The full hGA procedure was run 100 times on each of five folds from the calibration data set, producing in total 500 runs of the hGA. Then, the best solution from each run was summed, forming a feature rank vector. The rank of each feature was the number of times it was selected during the runs. The final feature selection was based on rank, with a limit of 0.05, i.e. if a feature was selected in more than 25 of the 500 runs, then that feature was included in the final hGA feature selection. Prior to the first BCI training session, the hGA was applied once on all data from the three calibration sessions (MI/Idle: 360 trials) and resulted in the selection of 118 and 110 features for subject 1 and 2 respectively (removing approximately 97% of all features). The same feature set was used throughout the whole BCI training intervention for training the classifier.

An SVM with a linear kernel (MATLAB v.2019, fitclinear) was used to classify the EEG features (reduced by the hGA) in real time. The classic hinge loss function was used to penalize predictions in the margin and appearing on the wrong side of the boundary. Ridge regression (or L2 regularization) was used to prevent large weights and dual coordinate descent was used to solve the dual optimization problem of the SVM, which is suitable when the number of features is high relative to the number of training trials. For each BCI training session, a new SVM model was trained on 100 randomly selected trials (50 MI + 50 Idle) from the calibration sessions and 32 trials (16 MI + 16 Idle) from the short calibration block of the current session, using the hGA features. This means that feature selection and SVM training were performed on partly the same data, however, the SVM testing/classification was performed on novel data sampled online during the BCI training. A single 250 ms window, centered at 1500 ms after task onset, was extracted from each training trial to be used when training the SVM. This time period has previously been shown to contain stable EEG activity related to MI (28). The SVM was tested on sliding 250 ms windows (step size 100 ms) starting 250 ms after BCI task onset until the end of each trial. Therefore, during the BCI training, the SVM model produced feedback every 100 ms by classifying the most recent 250 ms window of EEG data. The SVM output (i.e. prediction) determined the feedback provided to the patients (i.e. updated the hand position on the screen), meaning that feedback was provided approximately 110 times during a single BCI training trial.

## Adaptation of difficulty during BCI training

The threshold used to determine the feedback was modified in real-time to increase the difficulty of the BCI task for the patients. By default, the binary SVM classifies score (distance-to-bound) outputs above 0 as MI (generating positive feedback). However, this was modified to instead classify score outputs above the feedback threshold (FBT) as MI. Specifically, the FBT was chosen to provide 40% MI accuracy for each patient, such that on average 40% of classified EEG data windows during MI trials would provide positive feedback (open/close the hand, figure 4B). In general, this makes it more difficult for patients to receive positive feedback, but with approximately 110 feedback instances per trial, the patients are likely to receive positive feedback on almost all MI trials. To better understand how different levels of MI accuracy were experienced by the patients in this experiment, we illustrate in figure 2C four example trials with different classification accuracy. During MI trials, the hand on the screen starts in neutral position and moves towards its target position (fully opened or closed according to the task) with positive feedback. Low and medium-low single-trial accuracy (below 40 %) might produce some positive feedback, but the screen hand remains mostly near its initial state. With higher single-trial accuracy (above 40 %), there is a high chance of the hand reaching its target position and even maintaining its final position for a short time. The feedback threshold was initialized by performing a 5-fold cross validation on the SVM training data, yielding a score value for each calibration data window. The feedback threshold was initially set to the 60th percentile of the score value distribution of MI data windows. It was continually updated every 5 MI trials (using all available data) throughout the BCI training by adding new score values (based on the online MI data windows) to the MI score value distribution and recalculating the 60^th^ percentile. See figure 2B for visualizing the feedback update procedure.

**Figure 4.**
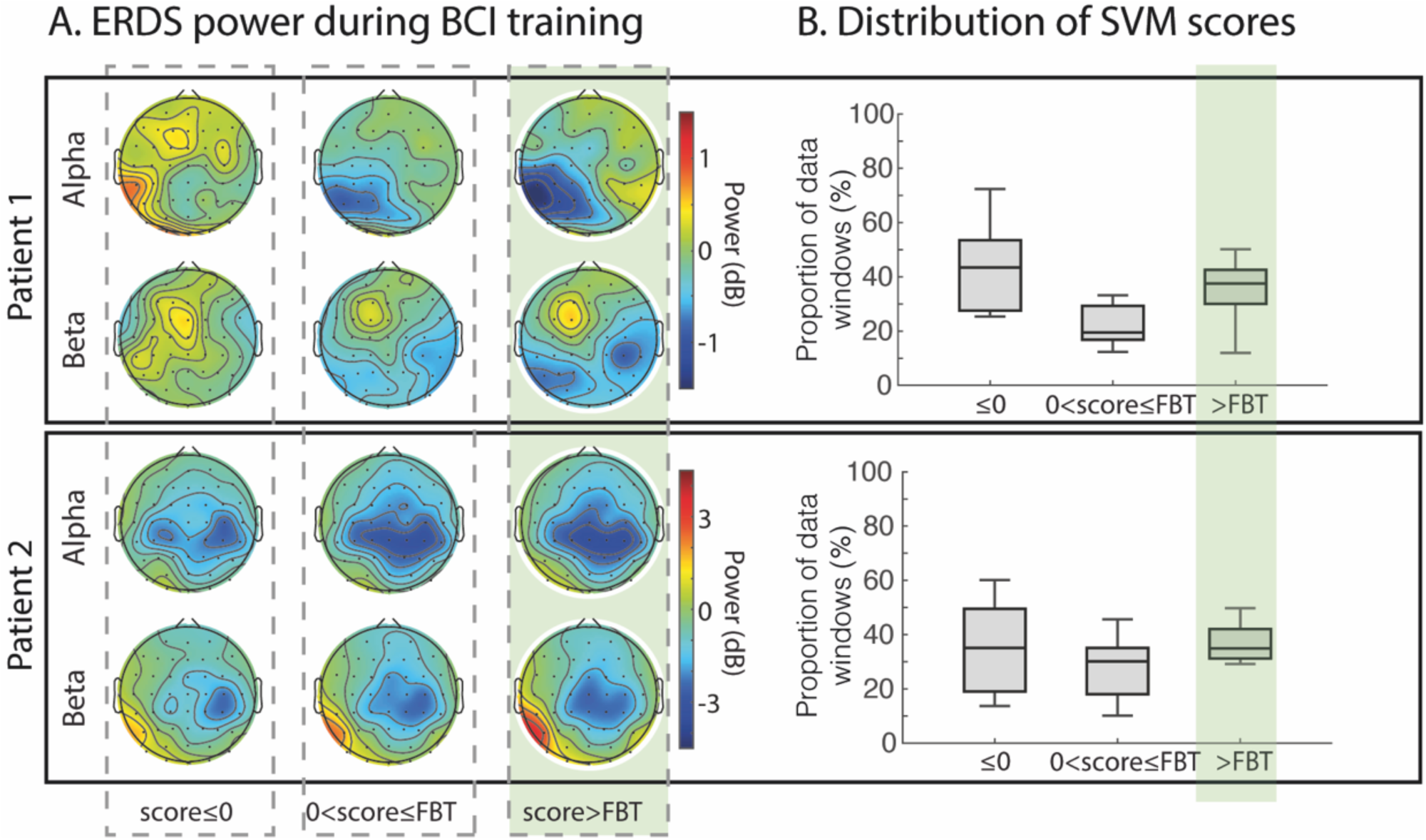
Distance-to-bound-related EEG features. A) ERDS power during BCI training. MI data windows from BCI training are categorized based on their SVM score (i.e. the output given by the classifier during the experiment) with respect to the feedback threshold (FBT). ERDS is calculated for each category in Alpha (8-12 Hz) and Beta (13-30 Hz). Column 1 illustrates MI data windows that received negative score, column 2 illustrates MI data windows that received positive score below the FBT, and column 3 illustrates MI data windows that received positive score above the FBT (i.e. positive feedback, visualized by the light green panel). B) Distribution of SVM scores. Boxes show the proportion of data windows, for each patient and across BCI sessions, for the different score categories in (A). Boxplot parameters are as follows: the line in the box correspond to the median, box edges show the first and third quartile, whiskers are drawn to the most extreme point within 1.5 times the inter-quartile range from the first or third quartile.

## Post-intervention offline analyses

In all offline analyses of the EEG data, the first second of each trial was removed since EEG often is highly non-stationary shortly after task onset. Power features from calibration and evaluation sessions were calculated using the same methods as were used during the online experiment (with overlapping windows and excluding the first second of EEG data). Online classification accuracy averages were recalculated post-experiment.

### Drift and distance metrics

The drift in the EEG signals was quantified by calculating the offset of the full SVM score distribution, including both MI and idle trial windows. The drift was defined as the peak of the distribution which was estimated by fitting a Gaussian (i.e. calculating the average score value). An illustration of the drift for two representative sessions of patient 2 is shown in figure 5A (i and ii). In addition to drift, the information content (in our case related to MI vs. idle) can be quantified by estimating the difference between the two SVM score distributions for MI and idle trial windows, respectively. In an ideal case with high information content, the two distributions would be largely separated. The difference between score distributions was estimated by calculating the distance between the peaks of the Gaussian fits. We illustrate the distance of a representative session of patient 2 in figure 5A (iii).

**Figure 5.**
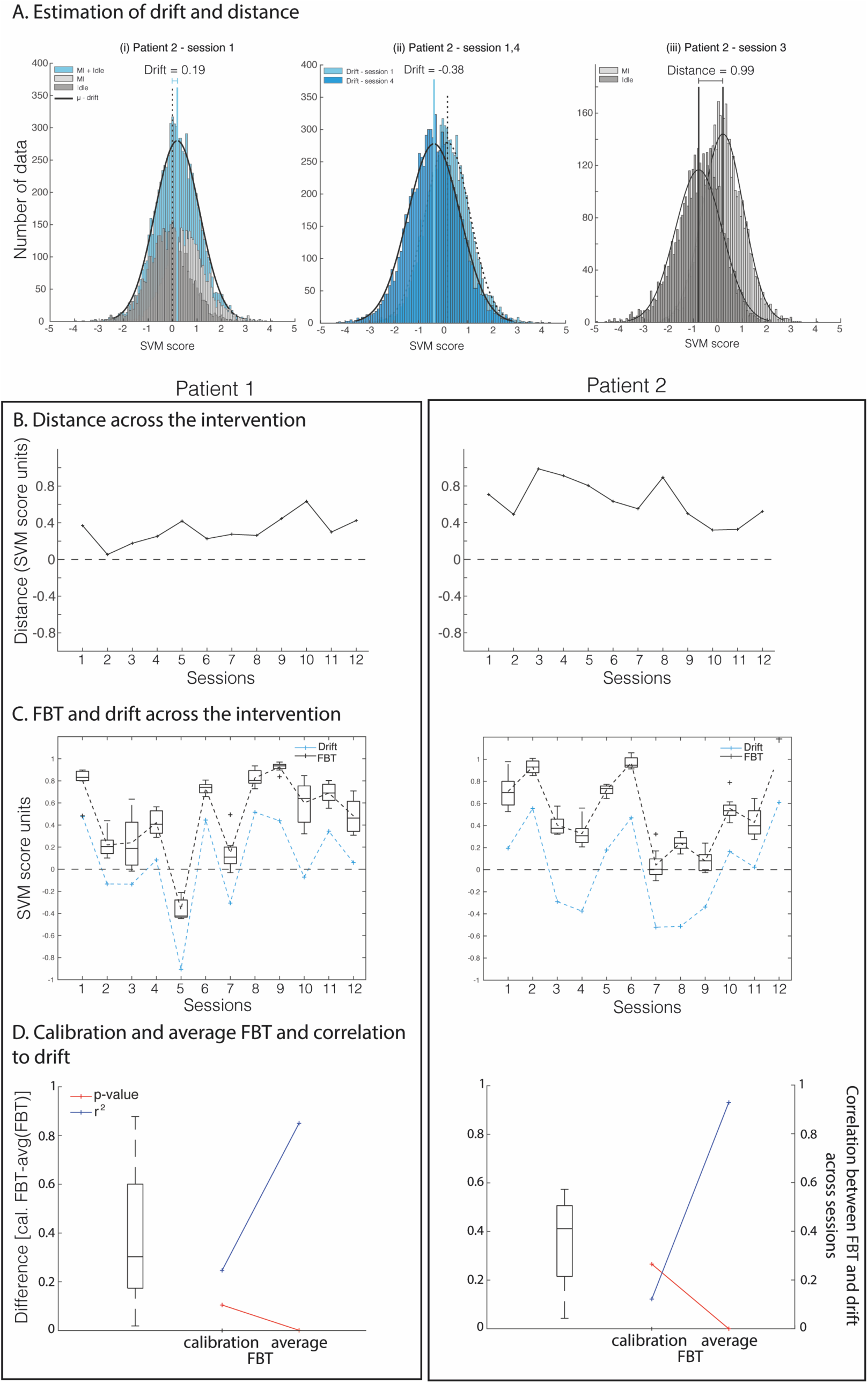
Distance, drift and feedback thresholds (FBT) across sessions. A) Estimation of drift and distance. SVM score distributions for representative patient 2 and sessions 1,3 and 4 are shown for MI+idle (blue), MI (light grey) and idle (dark grey) trials. The distributions were fitted with a Gaussian distribution. The drift was estimated by calculating the offset from zero to the peak of the Gaussian distribution of MI+idle trials. The distance was calculated as the difference between the peaks of the Gaussian distributions of MI and idle trials. B) Distance across the intervention. The distance is plotted for each session and patient. C) FBT and drift across the intervention. Boxes show the FBTs (n=8) for each session. The drift (blue) is plotted for each session. D) Calibration and average FBT and correlation to drift. For each patient, the box shows the difference between the calibration FBT and the first FBT calculated during each BCI training session. The lines correspond to the correlation metrics between session-by-session drift in (C) and either the calibration FBTs (for each session) or the within-session-averaged FBTs. Their correlation coefficients are shown in blue and the corresponding p-values are shown in red.

## Results

### SVM input features calculated by the hGA

To understand how the feature reduction with hGA in combination with the change in the feedback threshold affected the feedback given to the patients, we performed topographical analyses of EEG power features. The average weights of the SVM models across the 12 BCI training sessions allow us to visualize, for each frequency band, the hGA-selected spatial and frequential features, as well as their importance in the averaged SVM model during BCI training (figure 4A). In other words, the weight of each feature determines its relative contribution to the feedback provided to patients. The trained SVM model (boundary in feature space) consists of only a weight vector and a scalar. Classification is performed by computing the dot product between novel data (to be classified) and the weight vector and then adding the scalar. Without considering the change of FBT, the SVM model was defined such that a positive score value was classified as MI, while a negative score value was classified as idle. In practice, since all power values are positive, it means that positive weights represent ERS related to MI, and negative weights represent ERS related to idle. But the opposite is also true, that positive weights represent ERD related to idle, and negative weights represent ERD related to MI. This relationship allows us to infer the ERDS patterns during MI from the model weights in figure 3. To clarify, negative weights represent ERD for MI (blue) and positive weights represent ERS of MI (red). Several channels for both alpha and beta frequency bands across the sensorimotor cortical areas were selected by the hGA in both patients. These were partially overlapping but with different signs, indicating that the two patients had partly different ERDS patterns in alpha and beta related to MI. It is relevant to note that the selected features were bilateral in alpha for patient 1 and dominantly contralesional (and ipsilateral to the MI task) in both alpha and beta for patient 2.

### Adapting the difficulty of the MI-BCI training

We then wanted to explore the EEG power that led to positive feedback for the two patients separately. In other words, what were the EEG power patterns that were reinforced by positive feedback? To this end, we investigated the relationship between FBT-corrected score values (i.e. distance-to-bound) and neural engagement related to MI. More specifically, we analyzed the EEG power for different score values assigned by the SVM during BCI training. We specifically compared the contrast (MI – idle) for alpha and beta frequency bands in three categories based on the score value with respect to the current FBT (figure 4A). We decided to focus only on alpha and beta frequency bands since these are well-characterized in MI tasks (32). This analysis allows us to understand the underlying ERDS patterns as classified by the SVM models and that corresponded to positive feedback during the BCI training (green panel in figure 4A). The EEG power of all data windows in MI trials from the 12 BCI training sessions were categorized based on their SVM score value: i) lower than 0, ii) between 0 and the FBT, and iii) above the FBT. For this analysis (score categories) we excluded data of which the FBT was lower than 0, corresponding to approximately 7% of windows for each patient. Data in each category was then averaged and the mean idle activity across all windows and trials was subtracted from each MI category. The ERDS power topographies of these categories are presented in figure 4A.

Patient 1 shows a clear interaction between the intensity of the power and the score category. Stronger ERD can be observed over the contralateral sensorimotor cortex in alpha and bilateral sensorimotor cortex (slightly more posterior on the contralateral side) in beta. Stronger ERS can also be observed over the ipsilateral sensorimotor cortex in alpha. Patient 2 shows a slightly less obvious interaction effect, especially for ERDS power of score values above zero. Nevertheless, stronger ERD over the sensorimotor cortex, bilateral for alpha (including central locations) and dominantly ipsilateral for beta can be observed as score values are higher. An average of 38% and 35% of the data windows led to positive feedback to the patient 1 and 2, respectively (green panel in figure 4B), which is in line with how we predefined the experiment.

### The FBT accommodates drift between sessions

Next, we wanted to analyse how the FBT fluctuated session-to-session and whether drift and distance metrics changed in conjunction (see figure 5A for an illustration of how these metrics were calculated). First, we calculated the distance between MI and idle score distributions on a session-to-session basis to analyse how the information content, according to the SVM model, evolved during the intervention. From figure 5B, we show an upward trend of the distance for patient 1 while patient 2 displays a slight downward trend (Mann-Kendall’s test, patient 1: p=0.03, patient 2: p=0.06). Further, the drift and FBT both show high variability between sessions with no apparent trend for neither of the patients (figure 5C; Mann-Kendall’s test, patient 1: p=0.94 (drift) and p=0.45 (FBT), patient 2: p=0.95 (drift) and p=0.95 (FBT)). We excluded the initial FBT in figure 5C because it does not reflect the BCI training data (it was calculated only on calibration data, hereafter referred to as calibration FBT). There was a rather large decrease from the calibration FBT to the first update of the FBT which is shown in the box plot of figure 5D (patient 1: median=0.30, std=0.26, patient 2: median= 0.41, std=0.32). Furthermore, while we show a strong correlation between the average FBTs and drift for both patients (figure 5D; Spearman’s correlation, patient 1: r^2^=0.85, p=0, patient 2: r^2^=0.93, p=0), we did not observe any correlation between the calibration FBT and drift across sessions (figure 5D; Spearman’s correlation, patient 1: r^2^=0.25, p=0.10, patient 2: r^2^=0.12, p=0.27).

### The FBT decreases within sessions in conjunction with the drift

In addition to predefining the BCI-task difficulty by offsetting the FBT, we wanted to investigate how the online adaptation of the FBT within sessions behaved. First, in the way we designed the experiment, the FBT was updated based on all available data. As such, we included both the score values from the calibration data and from new incoming data windows from the BCI training. Because we included the scores of all data windows (of size 250ms) during the BCI trials (around 110 scores per trial), the data from the BCI training rapidly gained power in the update calculation of the FBT. We quantified the proportion of the new data with regards to all of the data that was used in the FBT update calculation (figure 6A). As expected, already for the first FBT update, the new BCI data represented a large majority of the total data amount (median of 89% for both patients). This proportion then decreased exponentially which is to be expected to reach better stability. Considering this, for both patients, we can observe larger fluctuation in the difference between the initial (calibration) FBT and the first FBT update across sessions (figure 6B; patient 1: std=0.38, patient 2: std=0.51). The variability of the difference between two consecutive FBTs then decreases until the second block (update #5), for which we observe an increase again followed by a decrease (figure 6B; update #5, patient: std=0.19, patient 2: std=0.14). We further investigated how distance, drift and FBTs changed within sessions (figure 6C). There was no significant trend for the distance within sessions for neither of the patients (Mann-Kendall’s test, patient 1: p=0.54, patient 2: p=0.11), although patient 2 showed a slight decreasing tendency. The drift significantly decreased within the session for patient 1 (Mann-Kendall’s test: p=0.019), whereas patient 2 only showed a slight decreasing tendency (Mann-Kendall’s test: p=0.17). The FBT decreased however significantly for both patients (Mann-Kendall’s test, patient 1: p=0.0020, patient 2: p= 0.00084).

**Figure 6.**
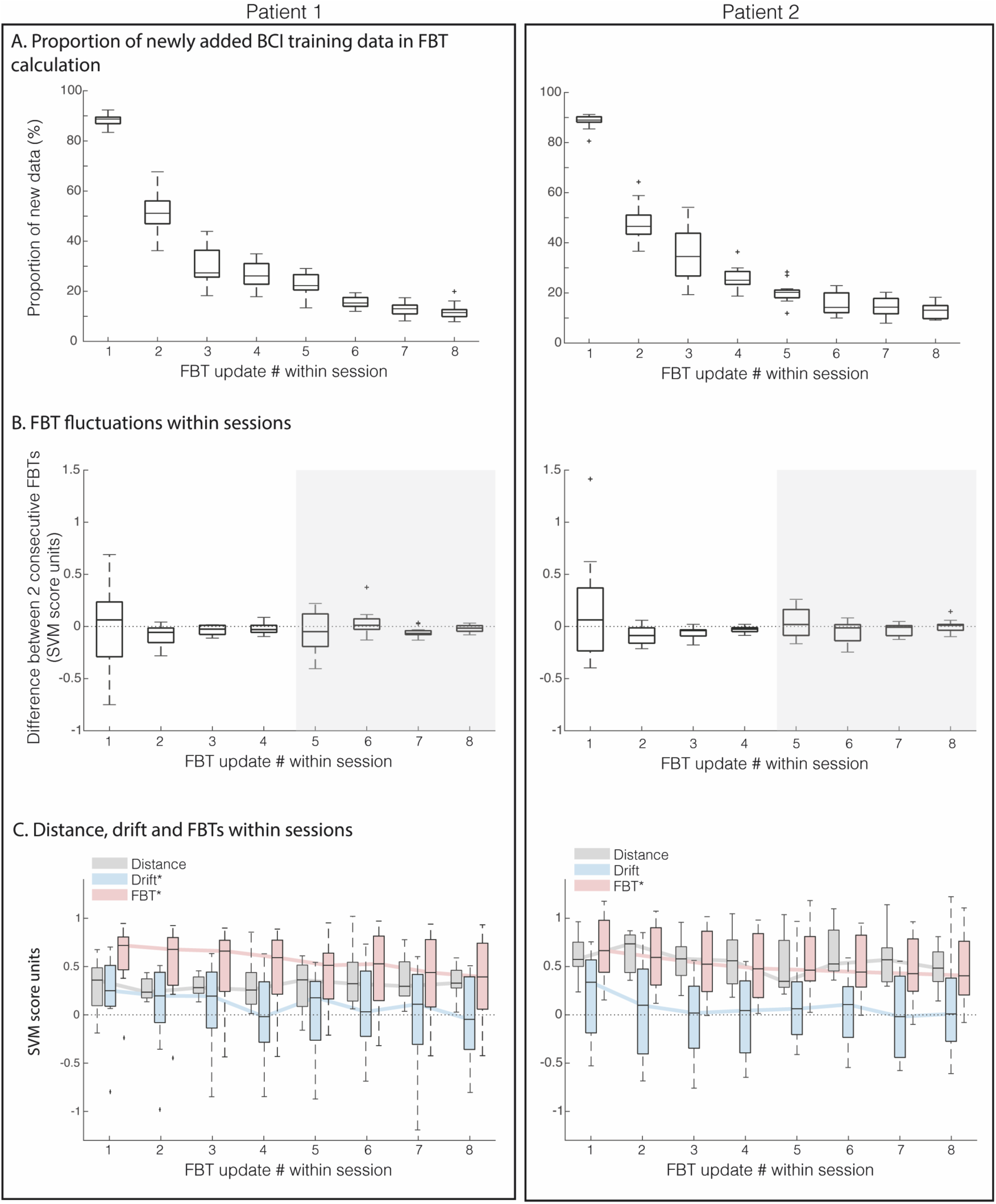
Distance, drift and feedback thresholds (FBT) within sessions. A) Proportion of BCI training data in FBT calculation. Boxes show, for each FBT update, the ratio of new data (BCI data) over the total amount of data used in the FBT calculation for all sessions (n=12). B) FBT fluctuations within sessions. Boxes show the difference between two consecutive FBTs for all sessions (n=12). First FBT update corresponds to the difference between the calibration FBT and the first updated FBT. The light grey transparent panel show the 2nd block. C) Distance, drift and FBTs within sessions. The distance (grey), the drift (blue) and the FBT (pink) are shown as boxes for each FBT update within the sessions. Significant trends at p<0.05 according to Mann-Kendall’s test are displayed with an asterix in the legends.

### EEG MI-related patterns and motor behaviour during BCI training

Despite the explorative nature of this study, we want to report on the evolution of MI-related EEG patterns, as well as the evolution of motor behavioural performance. First, focusing on the alpha and beta frequency bands, EEG power of (MI – idle) is plotted as a weekly average (figure 7A). Stronger alpha ERD over the contralateral sensorimotor cortex can be observed in patient 1, especially from the first to the second week of BCI training. Stronger beta ERD, dominantly over ipsilateral sensorimotor cortex can also be observed between the first and second week. The SVM distance, which reflects the overall discriminability between the EEG activity during MI versus idle trials, shows an increasing trend for patient 1 (figure 7B, statistical test in figure 5B). This trend is in line with what can visually be observed for the week-by-week alpha and beta ERD over the sensorimotor cortex. Patient 2 shows stronger alpha and beta ERD dominantly over the ipsilateral sensorimotor cortex in week 2. ERD then becomes weaker in week 3 and 4. These observations are also corroborated by the SVM distance showing a decreasing trend (figure 7B, statistical test in figure 5B).

**Figure 7.**
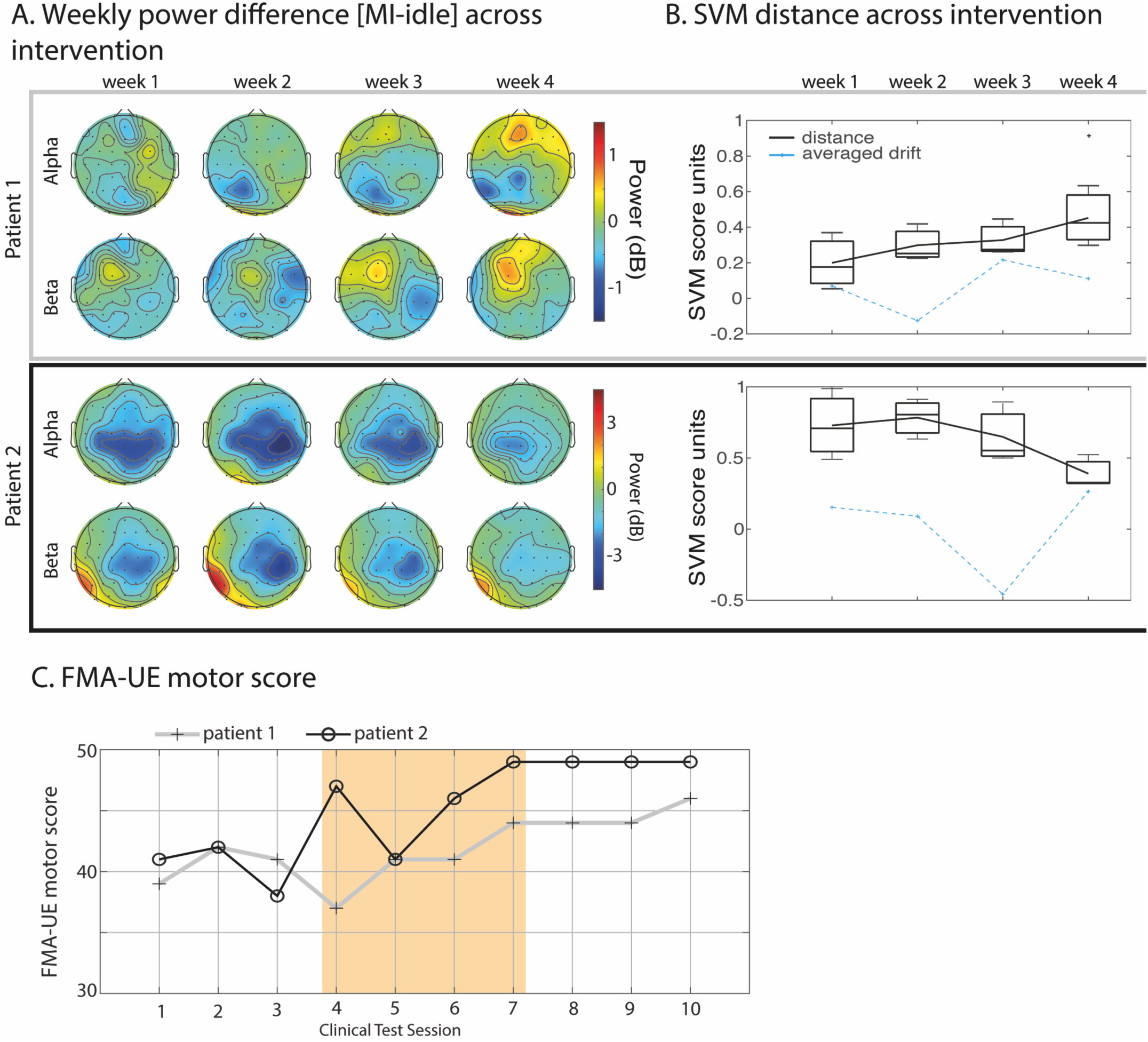
Evolution of MI-related activity during the intervention and correlation to motor behaviour. A) Weekly power difference [MI-idle] across intervention. Topographical plots of Alpha and Beta spectral power (MI – idle) averaged for each week of the BCI training. Positive power (red) signifies MI-ERS, i.e. that the EEG power is stronger during MI. Negative power (blue) indicates MI-ERD, i.e. that the EEG power is weaker during MI. B) SVM distance metrics across the intervention. Boxes show the distances for the three sessions each week. The drift averaged across sessions each week is shown in blue. Connecting lines are for visual purposes. Boxplot parameters are as follows: the line in the box correspond to the median, box edges show the first and third quartile, whiskers are drawn to the most extreme point within 1.5 times the inter-quartile range from the first or third quartile. Data points outside this range (outliers) are marked with a plus sign. C) FMA-UE motor score (patient 1-grey line with crosses, patient 2 – black line with open circles) for each clinical test session. The yellow-shaded area highlights the BCI training phase.

Motor-related behaviour evaluated with the FMA-UE scale shows clinical improvements for both patients. Patient 1 increased from 40.7 to 44.7 (+4) and patient 2 increased from 40.3 to 49.0 (+8.7) when comparing measurements before and after the BCI training (averaging calibration and evaluation data points i.e. before and after the yellow shaded area in figure 7C). Both subjects also showed a corresponding amount of change on the the FMA-UE hand subscale (7 to 12 and 9 to 11 points, not shown in figure). The Pearson correlation between weekly global FMA-UE motor scores during the BCI training (yellow shaded area in figure 7C) and median weekly distances revealed high rho-values, 0.95 and-0.80 for patients 1 and 2, with p-values 0.05 and 0.18 respectively.

## Discussion

This clinical case study has explored how the distance-to-bound approach can be used to adapt the difficulty in BCI training of two chronic stroke patients who participated in a 4-week MI BCI training intervention. Using a multiple-session design, we investigated the stability of this approach both between and within sessions. We further co-related its behaviour with the drift in signals both between and within sessions. Finally, we explored how it related to cortical modulations and behavioural outcomes of the two chronic stroke patients. The study adopted a within-patient explorative analysis approach and highlights the importance of this kind of analysis for personalization of BCI protocols.

### Patient-specific MI-related ERDS patterns during BCI training

For interpreting cortical activity in stroke patients, as they typically manifest differences in lesion size, location and thus brain activity, it becomes particularly important to consider patients individually. The two stroke patients in our study displayed differences in gender, age and lesion profiles. The heterogeneity of stroke patient profiles is a challenge in BCI research, and we strongly believe personalization is key in addressing this challenge. The use of machine learning in BCIs allows for building models on each patient’s data separately. In this study, we also applied an adaptive procedure during the BCI training, and we expect this approach to adapt to each patient separately. In our study, the two patients produced somewhat different ERDS patterns related to hand MI during the BCI training as seen in figure 4A (column 2-3) and figure 7A. The patterns partly overlap with similar hand MI EEG patterns that is typically observed in healthy individuals when simultaneously presenting online visual feedback (33), namely bilateral alpha,– and beta ERD above the sensorimotor cortex with dominant contralateral ERD and ipsilateral ERS in upper alpha and beta (17). During MI-BCI training, patient 1 produced contralateral ERD and weak ipsilateral ERS across parietal electrode locations in alpha as well as a somewhat asymmetric bilateral ERD across centroparietal locations and contralateral ERS across frontal locations in beta. However, the contralateral ERD (alpha and beta) for patient 1 was localized at parietal electrodes P(1,3,5), rather than the expected central C(1–6) (17,34) or centroparietal electrodes CP(1–6) (35). Meanwhile, ipsilateral beta-ERD is localized mainly at CP4, revealing an asymmetric mirroring along the coronal/frontal plane. This posterior shift of contralateral ERD might be explained by the location of the lesion. MRI images and EEG topographies in figure 1 reveal how the lesioned area of patient 1 affects a large part of the contralateral hemisphere. Electrodes P(1,3,5) are located at the edge of the lesioned area (the peri-lesional area), which could be a critical area for functional recovery (36,37). Patient 2 displayed a bilateral ERD in alpha during MI-BCI training, including the midline, symmetrically centered around centroparietal electrodes in both hemispheres (figure 4A and 7A). Beta ERD was also observed to be bilateral but with ipsilateral dominance. One possible reason for the more symmetric ERDS pattern in patient 2 could be that the lesion is frontally localized (at F3, see figure 1), and does not overlap with the sensorimotor area to a degree that would visibly affect the ERDS pattern. Thus, the ERDS pattern of patient 2 is more similar to those of healthy individuals’ (e.g. (38)).

Patient-specific analysis or grouping of similar patients (such as (39–41)) is crucial since averaged results may not represent individual stroke patients. Subject variability is a problem for generalized BCI classifiers (42) and lesions with varying location and size can cause further deviation from typical ERDS patterns (43,44). Multivariate pattern analysis (e.g. machine learning) allows for better subject-specific modelling, but introduces the risk of modelling physiological noise rather than true brain signals (45), especially if the feature space is greater than the training data set (12). To reduce the risk of incorrect modelling, we compare each patient’s MI pattern with typical MI patterns observed in healthy individuals while also considering the location and extent of the lesion. While both patients have near-typical MI-related ERDS patterns in alpha and beta, their activity patterns differ as well as their lesion profile and hence patient averages would not provide useful information. Instead, it would attenuate the effects and lead to biased interpretations.

Rehabilitative BCIs are based on promoting neuroplasticity through neurofeedback that reflects motor activity of a patient’s brain (1). Therefore, motor-irrelevant feedback is unlikely to promote clinical improvements, similar to how sham-feedback has been observed to fail in producing clinical improvements in BCI rehabilitation (46). More detailed analyses of patient-specific ERDS patterns related to MI and similarity to typical MI-ERDS patterns in healthy people (such as in (40,41)) can lead to valuable insights for the development of personalized BCI rehabilitation, increasing the effectiveness by guiding the use of machine learning and multivariate pattern analysis.

### Encouraging stronger MI-related EEG activity by adjusting the FBT

The difficulty of controlling a BCI lies in the ability to generate the EEG activity patterns that produce positive feedback (i.e. moves the hand in the intended direction). By changing the FBT, we can set the strength of the EEG power patterns that are required of the patient to generate positive feedback. This level of required strength was determined from each patient’s own EEG power during MI trials by setting the FBT to the 60th percentile of the individual MI score distribution. In figure 4A, we show the relative strength of EEG power that was required for receiving positive feedback in our BCI task (green panel). This power can be compared to the EEG power that would have been required in a standard BCI task (i.e. without increasing the difficulty) by comparing the topographical plots of *0<score<FBT* and *score>FBT* in figure 4A. As is visible in the figure, the ERD of alpha and beta across sensorimotor regions is higher for *scores>FBT* (i.e. for positive feedback) for both patients, showing that patients needed to induce stronger MI-related activity (16,17) to receive positive feedback. As we assume that producing stronger brain activation (more discriminable from idle activity) is more difficult, we thus show that we have effectively changed the difficulty of the BCI task.

We further observed that the FBT slowly decreased throughout each session for both patients (figure 6C). One might interpret this result as the MI-related EEG activity becoming weaker (evolving towards the idle-related activity) as a session progressed. However, judging from the distance measures between MI and idle trials remaining at the same level (figure 6C), this rather suggests that the EEG signals were slowly drifting within sessions. This is further corroborated by the decreasing drift in figure 6C. Our aim was to encourage stronger MI-related activity during the BCI training, and indeed, positive feedback represented increased ERDS power in the alpha and beta band (figure 4A). However, it might have been too difficult for the two patients to improve (i.e. increasing the threshold) within a session. Many BCI studies suggest that at least 70% or higher BCI classification accuracy is necessary for feeling in control when using a BCI (47). However, the need for high BCI performance is based on the utility of BCIs as assistive tools, such as speller BCIs. This is very different from the rehabilitation user-case. Neurorehabilitation after stroke is founded on activity-based neuroplasticity (6), which is based on practicing a motor task that requires neural adaptation to succeed. In other words, the level of difficulty in performing a task represents the amount of neural adaptation required to complete the task. For rehabilitation, it can be beneficial to successively increase the difficulty or complexity as the patient gains mastery of the task (48). This is consistent with best practice recommendations for physical stroke therapy, suggesting repeated every-day tasks with increasing difficulty or complexity (49). The pre-defined difficulty level in this study might have been set too high and further research is needed to better understand what an optimal difficulty level might be. However, we contribute with a conceptual study on how the distance-to-bound can be used to adapt the BCI difficulty.

### The online adaptation of FBT accommodates signal drift

There is ample evidence that changing context from a no-feedback to a feedback condition leads to modulations in EEG activity (50). This phenomenon can also be observed in our data by the larger differences between the calibration FBT and first FBT of each block (i.e. update #1 and #5 in figure 6B), indicating a drift of the EEG signals from calibration (i.e. no feedback) to BCI training. This poses limitations on BCIs driven by machine learning since they require data for calibrating or building an initial model before neurofeedback can be provided to users. An additional limitation is the non-stationarity of EEG signals across sessions, blocks and sometimes even within trials. To overcome these limitations, this study opted for an adaptive SVM classifier trained on data both from prior calibration sessions and from a small calibration block in the current BCI training session. This calibration data set consisted of 130 data samples, one sample per trial, as input to build the SVM model and calculate an initial FBT. During the BCI training, since SVM score values were calculated from data in windows of 250ms and all scores were used to update the FBT, this feedback-related data rapidly outgrew the size of the calibration data (figure 6A). We show that this approach resulted in an increasing stability of the FBT within sessions (i.e. differences between consecutive FBTs converged to zero; figure. 6B). A potential limitation of the frequent update of the FBT is the impact of fluctuations in the MI-related signal due to fatigue, attentional lapses or other fluctuations. The way we defined the FBT update procedure, the impact of new data (with potential fluctuations due to fatigue etc.) reduces exponentially (see figure 6A). This makes sense as fatigue-related fluctuations are hypothetically more frequent during the second part of the session and will therefore, in our design, have less impact on the updated FBT. This will however need to be tested in a larger sample of participants.

The online adaptation of the FBT, independent of its predefined level, was further shown to accommodate class-independent drift in the EEG signals, both within and between sessions (figure 5 and 6). The high correlation between the FBT and the drift indicates that even though the FBT was calculated from score values only from MI trials, this method accounted well for idle-related drift as well. Further, we showed that the distance did not correlate with drift, indicating that the MI-related information evolved as an independent process.

### Within-patient analysis of motor behavioural outcomes and the correlation to the strength of MI ERDS patterns

BCI performance is generally measured as the rate of correctly classified instances (12) with high performance being crucial for controlling a device. However, rehabilitative BCIs are intended to facilitate neural recovery and achieve clinically meaningful improvements in behaviour (51). Although high classification accuracy generally implies significant decodable information in the brain activity, performance of rehabilitative BCIs cannot be evaluated solely using this measure. Instead, it is often evaluated with standardized clinical tests, such as FMA-UE motor score which is commonly used to measure changes in motor function post rehabilitation. This study was limited by only having two patients, however it has aimed to provide value by thoroughly monitor and analyse the interaction between behavioural, neurophysiological and computational factors for each patient. It capitalizes on the multi-session nature of the experiment which is less common in BCI research. Considering the specific patient group studied here, personalization will be necessary in future BCIs. Therefore, the increased practice of intra-subject analyses will be required to understand differences and optimize BCIs. An intra-subject approach gains in statistical power and could better accommodate for the between-subjects variability (well-documented in EEG-research (42,52–54)) of activation patterns and brain plasticity specifically observed in stroke patients (18). For this approach, evolution of brain activity throughout the rehabilitation intervention as well as feedback-related (specific to the classification model) brain activity should be described and correlated to clinical test outcomes, stroke lesion profiles, symptoms and other measurement modalities to ensure BCI-specific outcomes.

Throughout the intervention, both patients improved on the FMA-UE (figure 7C). CID for chronic stroke patients with moderate hemiparesis ranges from 4.25 to 7.25 FMA-UE motor score (25). CID on the FMA-UE scale was reached at large by patient 2 (+8.7), whereas patient 1 came close (+4). The across-patient average improvement in this study was +6.35 FMA-UE, which is at the high end of the CID range and can be compared with two larger stroke-BCI studies who achieve +4.5 (55) and +3,4 (56). One could argue that the motor improvement observed for both patients can be attributed to the clinical tests performed once a week during the intervention and/or to the attempted ME trials performed in the beginning of each BCI session, as these tests and trials resemble physiotherapy. Furthermore, physiotherapy in combination with MI may produce clinical improvements even without BCI feedback (57). However, if the clinical tests or the attempted ME trials caused motor improvement then we would expect to see an improvement during the pre-and post BCI training sessions. When inspecting the pre-and post-training sessions in figure 7C (test point 1-3 and 7-10) we found no sign of improvement. The FMA-UE is stable at these times and Patient 1 decreased in FMA-UE motor score over test points 2 to 4 (figure 7C), indicating that the clinical motor tests and attempted ME performed prior, during and post BCI training had little or no positive clinical effects.

Further, we wanted to investigate how the MI EEG-pattern changed over the four weeks of BCI training by analysing topographies of weekly averaged ERDS power in the alpha and beta bands (figure 7A). Specifically, we were interested in whether the patients’ EEG activity had evolved towards that of the target activity reflected in the positive feedback (represented by the green panel in figure 4A). An overall comparison between the weekly and target ERDS patterns shows that the magnitude of the targeted ERDS pattern varied between different weeks. Specifically, patient 1 had weaker MI pattern during the first week of BCI training which became stronger during the following three weeks. Patient 2 had stronger MI pattern during the second BCI training week, followed by a decreasing magnitude during the last 2 weeks (figure 7A, B). These changes in alpha and beta power are corroborated by the week-to-week distance measure (figure 7B), suggesting that MI-related information, as defined by the SVM model, increased across the intervention for patient 1 and decreased for patient 2. The high correlation and anticorrelation between the distance and the FMA-UE score during BCI training, for patient 1 and 2, respectively, demonstrates the importance of correctly configuring a machine learning model for providing functional feedback. Both patients improved their motor function in a clinically meaningful way, however the observed modulations of MI-related EEG activity patterns were not as expected for patient 2. Our interpretation is that the feedback provided to patient 2 was not optimal for enhancing motor function, however, the intensive practice of MI (albeit with non-optimal feedback) still led to behavioural improvements in motor function. As this result does not in any way invalidate our results with regards to the distance-to-bound approach in adapting the difficulty and accommodating for signal drift, it does provide us with additional insight of the complexity in determining functionally relevant feedback after stroke.

Activity measured with fMRI has been observed to typically increase in both ipsi– and contralesional motor areas in the acute phase of stroke (58). However, this seems to be a temporary phenomenon in patients with good motor recovery (59). In other patients, for which the overactivity of the contralesional hemisphere remains, motor deficits usually persist (59,60), suggesting that it has detrimental effects on motor recovery. Specifically for EEG, increase of the ipsilesional in conjunction with decrease of the contralesional sensorimotor rhythm (SMR) has been observed to correlate with functional recovery after stroke, whereas enhanced contralesional SMR was observed in one non-recovered patient (40). The functional relevance of activity changes post-stroke remains under debate, and the mechanisms enabling plasticity and functional recovery are not well understood (61). However, considering what we know so far, it is surprising that BCI protocols do not take this into account in the design of neurofeedback. In this study we show a striking case of motor functional gain related to more focal and weaker ERD activity across sensorimotor regions after BCI training. Due to the small sample size in this study, we believe that an in-depth discussion on the mechanisms enabling functional recovery post-stroke would be out-of-the scope. For this, we refer to other targeted studies (e.g. (61)). Our results emphasize the importance of patient-specific analysis and deeper investigation into how EEG activity is expected to evolve as a patient recovers.

### Challenging the use of BCI illiteracy as inclusion criteria in rehabilitation

Relevant feedback is crucial for successful recovery when training with a BCI (56) and high classification accuracy has been linked to better clinical improvements in BCI rehabilitation (57,62). This case study is in line with this finding as patient 2 showed larger distance between MI and idle SVM score distributions (correlating to higher classification accuracy, see supplementary figure 4) during BCI training and hence increased more in FMA-UE motor score as compared to patient 1. However, we note that patient 1 reached close to a clinically meaningful improvement in motor behaviour, despite having near chance-level BCI performance throughout the BCI training phase (average classification accuracy of 55.2%, see supplementary figure 4). Interestingly, we found that the BCI provided MI-relevant neurofeedback to patient 1 despite chance-level BCI performance (figure 4A). It seems the low classification accuracy was not based on the BCI’s inability to detect the MI-related activity in patient 1, but the patient’s ability to consistently produce it. This is an important finding that adds to the debate on BCI illiteracy (63,64). Research suggests that approximately 30% of people may not be able to proficiently control a BCI (47), an inability often termed “BCI illiteracy” or rather “BCI inefficiency”. As proficiency is often defined as achieving at least 70% classification accuracy (47), our study opens the possibility for more inclusive BCI designs and the possibility of chance-level BCI users to benefit from MI-BCI rehabilitation. Due to screening procedures, labelling patients as BCI illiterates occasionally excludes them from participating in BCI training (55,65,66) even though it may in fact benefit them (63). Despite the claims of requiring high BCI performance, it has been shown that low classification accuracy (60-75%) can be sufficient for clinical improvements with MI-BCI training (67). Our results suggest that it may be possible for extremely poor BCI-performing patients to benefit from BCI-based stroke rehabilitation, if the BCI classification model can promote functionally relevant brain activity which guides neuroplasticity towards recovery. Future work should be inclusive in the recruitment of study participants and investigate the potential of the distance-to-bound approach for adapting the difficulty in a randomized controlled trial of at least 10 participants per group.

Another important consideration is that in our study, the feedback reflected the joint EEG activity of both open and close MI. The classification of same-hand MI has been shown challenging, and the prediction is associated with a higher degree of uncertainty (68). Further, the underlying EEG activity is highly overlapping (38), which led us to rely on the instruction given to patients to perform the MI of the specific hand movement. However, this needs to be considered as the two MI tasks might not lie on the same difficulty level.

### The use of pre-selected features by mRMR reduction in MI BCI training after stroke

In this study, we pre-selected features based on a hierarchical genetic algorithm (hGA) to manage the high dimension of input features to the SVM classifier that can otherwise lead to overfitting. Training samples are inherently limited since they need to be collected for each patient separately, however, SVMs generally show good performance for high feature dimension in combination with small training sets (69). Another potential limitation is that GA:s are susceptible for ending up in local minima. To overcome this, we ran the algorithm 500 times on different training data folds and randomly initialized populations, and extracted features based on rank (70). Additionally, we have carefully examined the features selected by the hGA (figure 3) and discuss how they relate to the observed ERDS patterns during the intervention.

The ERDS power patterns for our two patients partially overlapped with the selected hGA features. In our study, the feature selection was performed using an hGA with mRMR as reward function. mRMR rewards relevance (correlation with task) while penalizing redundancy (correlation with other features) (31) which may lead to a discrepancy between the pattern of selected hGA features, and the patients’ MI EEG ERDS features. Additionally, it is also important to note that the hGA selection is performed on the calibration sessions (i.e. without feedback). The general observation for both patients is that the ERDS power spans a wider area as compared to the hGA, which is reasonable considering the theoretical basis of the hGA algorithm. The use of mRMR may also have possible consequences for brain plasticity. As such, any feature, despite correlating well with MI, may be omitted in favour of a similar feature. For example, in figure 4A we observe that the MI pattern of patient 1 corresponds to contralateral alpha-ERD in multiple channels: C-(1,3,5), CP-(1,3,5) and P-(z,1,3). However, only CP5 was selected for classification (figure 3). It seems that the entire contralateral alpha-ERD area was well represented by CP5 alone, and other similar channels were penalized as redundant. This may be a negative trait for stroke rehabilitation, since the most discriminatory frequency components may vary from session to session (42,71), and functional recovery in stroke patients may be accompanied by evolving EEG patterns (72). If this was the case, then mRMR-based feature selection may have counteracted the BCI training in this study since the feature selection was performed only on the calibration sessions and was not updated throughout the intervention. Hypothetically, a full re-train of the model where all features are considered could enable finding the session-optimal pattern for neurofeedback, although adding the drawback of requiring new data for the training of the model for each session.

## Data Availability

Due to the small data sample (n=2) and strict ethical considerations, data cannot be made available.

## Conclusion

In this study, the feasibility of the distance-to-bound approach to manipulate difficulty during BCI training was explored. Our results suggest that although the difficulty was set very high, patients received positive feedback mainly for strong alpha and beta ERD across sensorimotor areas, thus reinforcing them to produce this activity. We show, with a multi-session experimental design, that adapting the FBT online accommodated class-independent drift in EEG signals both between and within sessions. We further show that our adaptive strategy resulted in the FBT stabilizing within each session. Further, during the four-week BCI training intervention, we observe modulations in the MI-related EEG power, which was correlated and anticorrelated to improvements in motor function for patient 1 and 2, respectively. These results showcase the necessity of tailoring the feedback to each patient depending on stroke lesion and hence EEG activation profile. We believe these results provide valuable insights that can guide future research on BCI-based stroke rehabilitation towards more effective methods and training protocols. It is important to emphasize that this study is explorative in its nature and cautious interpretations of the results are needed.

## Declarations

### Ethics approval

The study has been conducted in accordance with the principles embodied in the Declaration of Helsinki and complied with local rules and regulations according to the Swedish Ethics Review Authority (dnr. 2019-01577). Both participants signed a written consent form before participating in this study.

The study was registered as a clinical trial at clinicaltrials.gov (NCT03994042).

Consent for publication was given by all authors.

### Availability of data and materials

Due to the small sample size (n=2) and strict ethical considerations, restrictions on data sharing apply.

### Authors contributions

Conceptualization, EA and JT; Methodology, EA, JT, JP and SP; Patient recruitment, SP; EEG data collection, JT; MRI data collection, JT, JP and SP; Clinical data collection, SP; EEG data analysis, EA, JT and MJA; MRI analysis, JP; Writing – original draft, EA and JT; Writing – review and editing, EA, JT, MJA, JP and SP; Funding acquisition, EA; Supervision, EA.

## Acknowledgement

We thank Prof. Jörgen Borg, MD, for help in the recruitment of patients and constructive discussions during the project’s planning and design.

## Abbreviations

MI: Motor Imagery
ME: motor execution
EEG: Electroencephalography
BCI: Brain-computer interface
MI-BCI: Motor imagery-based brain-computer interface
MRI: Magnetic resonance imaging
FMA-UE: Fugl-Meyer assessment for upper extremity
SVM: Support vector machine
EMG: Electromyography
EOG: Electrooculography
hGA: hierarchical genetic algorithm
mRMR: Minimum redundancy maximum relevance
ERDS: Event-related desynchronization and synchronization
ERD: Event-related desynchronization
ERS: Event-related synchronization
FBT: Feedback threshold

## Footnotes

1 The absolute value was taken to avoid cancellation due to possible directly proportional and inversely proportional relation between features or between features and classes.

