## Supplementary materials for "Exploration of using “distance-to-bound” to manipulate the difficulty during motor imagery BCI training after stroke – A clinical two-cases study"

### Inclusion and exclusion criteria of recruited patients

The patients fulfilled the following inclusion criteria: age > 18 years; < 6 months since first time stroke onset and with remaining hemiparesis in upper extremity; able to participate fully in the intervention including screening of cognitive function with the Cambridge Neuropsychological Test Automated Battery (CANTAB) (1–3); able to perform Functional Magnetic Resonance Imaging (fMRI); able to passively extend the wrist 15 degrees and extend fingers fully with a neutral position of the wrist. In addition, need to be able to voluntarily control the power of their grip when requested according to the Visuomotor force tracking method (4) and/or according to the clinical assessment of a therapist (while holding the patient´s hand). According to the FMA-UE scale (5) participants should accomplish <14 points on the hand subscale (C) in addition to < 48 points on the total motor score (equivalent to moderate disability in the upper extremity (6).

Exclusion criteria were other neurological or musculoskeletal disease/injury, contagious disease or treatment with botulinum toxin in the upper extremity during the past 3 months, current or history of epilepsy, hearing problems, metal implants in the brain/skull cochlear implants, any implanted neurostimulator, cardiac pacemaker or cardiac implants of metal, infusion device, any other neurological disorder, pregnancy, current or history of severe psychiatric disorder with need for pharmacological treatment (7).

### The intervention outline

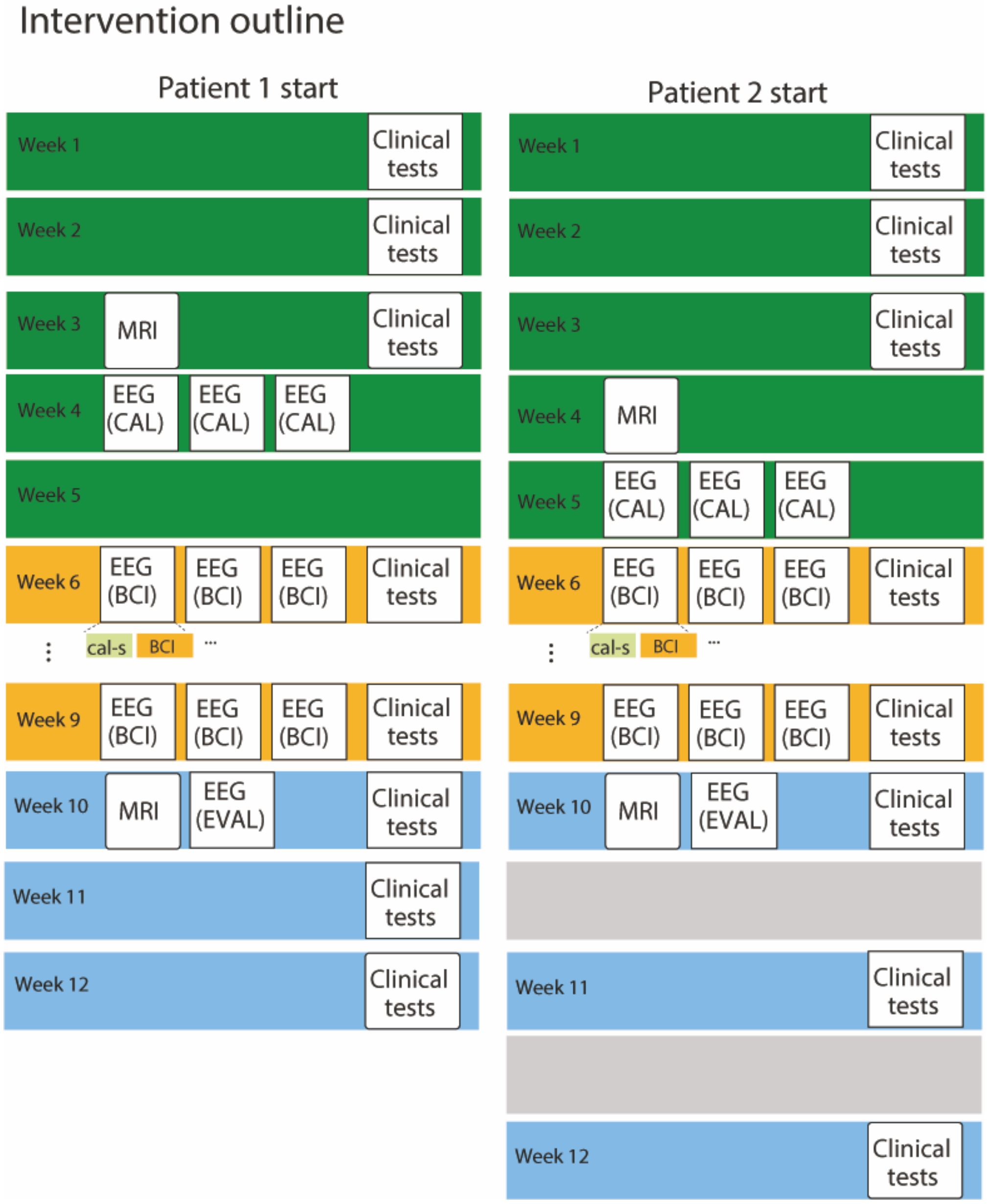


Supplementary Figure 1. **The intervention outline**. Each bar represent 1 consecutive week, and white boxes explain what activities were performed by the patient. The intervention consisted of three phases: calibration (dark green), BCI training (yellow), and evaluation (blue). EEG (CAL) and EEG (EVAL) are EEG recordings without feedback, while EEG (BCI) consists of a shortened calibration session followed by a BCI training sessions with real-time feedback. Grey bars represent weeks where the patient was unavailable for recording, thus delaying subsequent tests.

### EMG and Accelerometer data

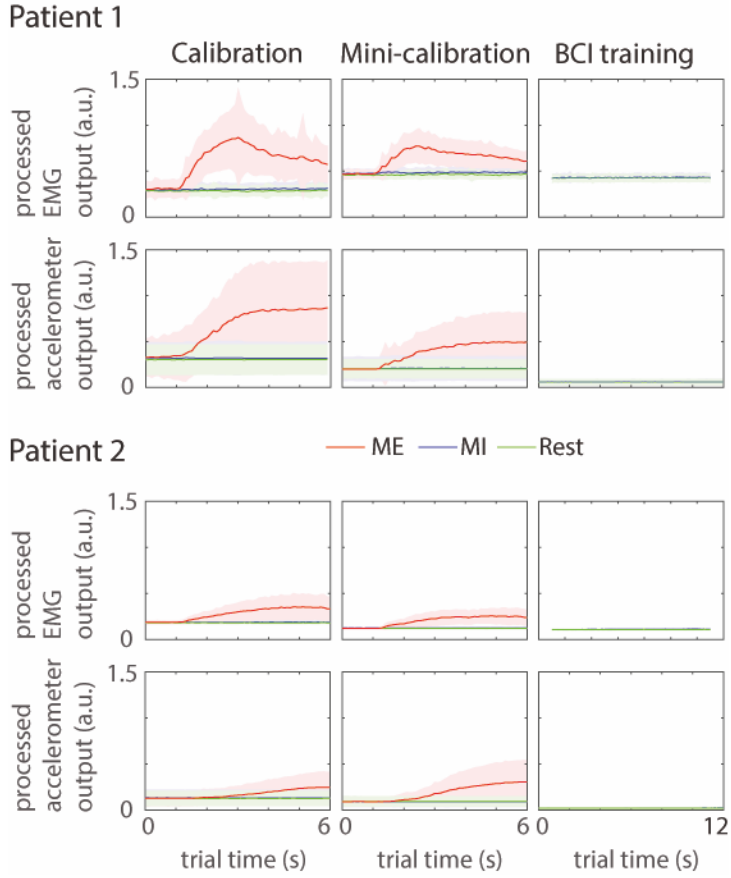


Supplementary Figure 2. **EMG and Accelerometer data.** A) EMG data for each task (MI, Rest, ME) averaged across trials and sessions. EMG data is highpass filtered at 5 Hz (order 3000) and bandstop filtered at 45-55 Hz (order 3000). Absolute values of EMG data were smoothed (moving mean, 100 samples) and downsampled 1:100 (selecting every 1 hundredth sample in a trial). The mean trial from all sessions of each type (calibration, short calibration, BCI training) was scaled by ME in calibration and presented with ste (shaded area). B) Accelerometer data averaged across trials and sessions. Accelerometer data was subtracted with its session-mean (no filtering). Absolute values of accelerometer data were smoothed (moving mean, 100 samples) and downsampled 1:100 (selecting every 1 hundredth sample in a trial). The mean trial from all sessions of each type (calibration, short calibration, BCI training) was scaled by ME in calibration and presented with ste (shaded area).

### Center of ERD peak calculations

To quantify the spatial change in EEG power prior versus post BCI training, center of mass equation (equation 1) was adapted to the EEG data in order to highlight the center of an event-related desynchronization (ERD) field (equation 2). In equation 1, R is the center of mass, $m_i$ is the i'th body's mass and $r_i$ is the i'th position in space. Two changes were made to this equation to produce center of ERD. First, the mass is replaced with the relative ERD calculated from average (MI – Idle) in each channel and frequency band. Second, the ERD is raised to the scaling factor p. The relative (MI – Idle) power in a channel and frequency band (band feature) corresponds to an object with mass, and position vectors were extracted from the topography. Only ERD was considered in these calculations, i.e. channels with event-related synchronization (ERS) were omitted.

1. Center of Mass $R = \frac{\sum_{i=1}^{n} m_i r_i}{\sum_{i=1}^{n} m_i}$
2. Center of ERD $R = \frac{\sum_{i=1}^{n} erd_i^p r_i}{\sum_{i=1}^{n} erd_i}$

The ERD scaling factor, p, was set to 5 as it was observed to better emphasize the true center of the ERD field. Since the ERD do not vary substantially between channels, a lower factor p causes the center to be placed near the midpoint of the brain due to the high number of channels influencing the formula. Note that the scaling factor is not used for the topography plot in figure 7, only for calculating center of ERD.

### No changes in MI-relevant EEG patterns post-intervention

To investigate the effects of the BCI training on MI activity without feedback, the ERDS and MI pattern strength was compared before (3 calibration sessions) and after BCI training (1 evaluation session).

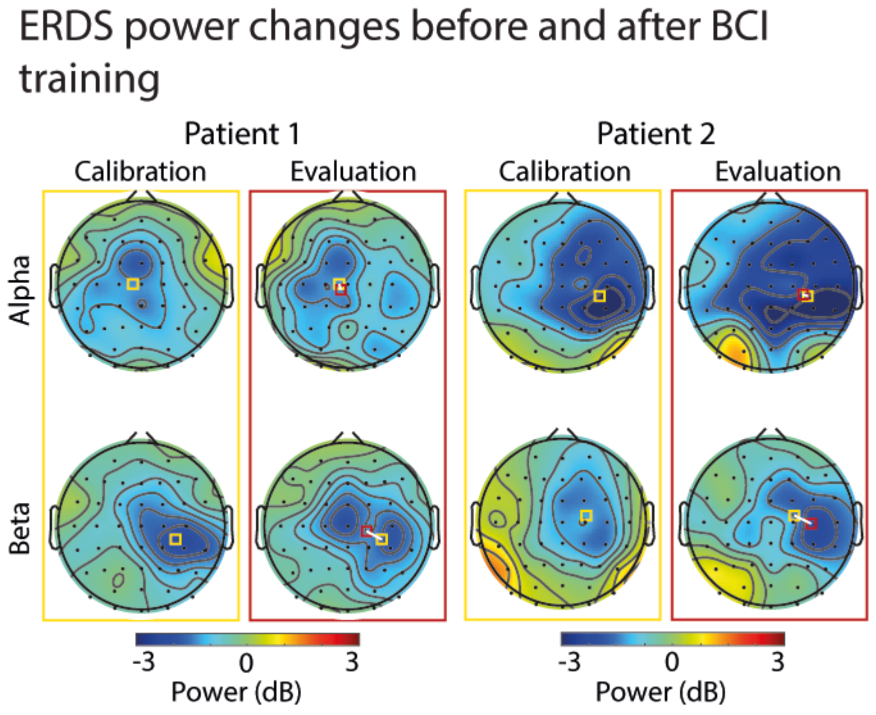


Supplementary figure 3. **MI-related activity before and after BCI training.** Topographies of ERDS during the calibration phase (yellow border) and the evaluation phase (red border) for alpha (upper row) and beta (lower row) frequency bands. The center of ERD power during calibration is represented by a yellow square and during evaluation by a red square. The evaluation topographies show both squares joined by a white line to emphasize the change of location of the centers of ERD prior to post BCI training.

Topographical data was calculated individually for each calibration session and then averaged for comparison with the evaluation session. The BCI training had negligible effect on ERDS patterns recorded without feedback, as seen when comparing calibration and evaluation data (supplementary figure 3). The center of ERD peak reveal that after the BCI training phase, patient 1 had extended the beta-ERD towards the contralateral side (electrodes C1, Cz, CPz) while patient 2 had instead a stronger beta-ERD more towards the ipsilateral side (C4, FC4, CP4). No change in alpha-ERD was detected for neither of the two patients.

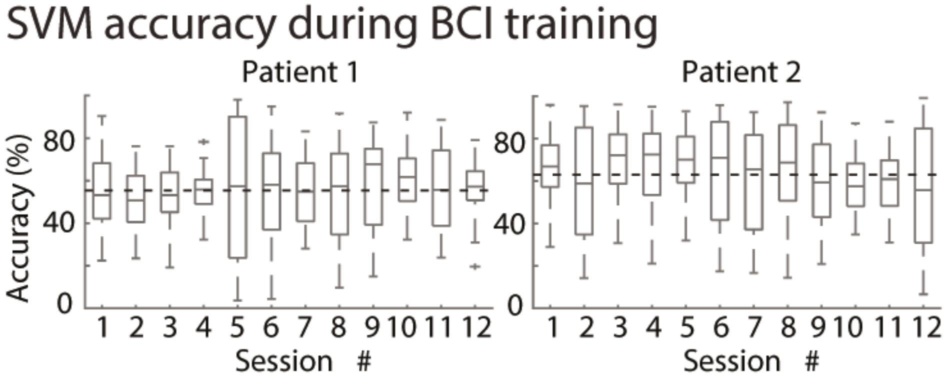


Supplementary figure 4. **BCI performance**. BCI performance is calculated as the unbiased (ignoring FBT) classification accuracy of both MI and idle trials. Each box represents trial-by-trial BCI performance in one BCI session. Boxplot parameters are as follows: the line in the box correspond to the median, box edges show the first and third quartile, whiskers are drawn to the most extreme point within 1.5 times the inter-quartile range from the first or third quartile. Data points outside this range (outliers) are marked with a plus sign.

SVM accuracy during BCI training

To evaluate how well the classification algorithm discriminated between MI and idle, we present the SVM classification accuracy of both MI and Idle during the BCI training with the feedback threshold set to zero (i.e. unbiased, supplementary figure 4). The unbiased classification accuracy of all (both) classes is the most common method for measuring BCI performance (8) and is more suitable for significance testing. Patient 1 achieved on average 55.4 % classification accuracy across the BCI training sessions, while patient 2 achieved on average 64.0 % classification accuracy. Using the look-up table from (9), with two classes and 960 trials (80 per session, 12 sessions), a classification rate above 55.2% is significant at alpha = 0.01 so both patients achieved significant classification accuracy. The look-up table has no entry for cases with more than 500 trials, so this entry was used (entries with fewer test samples yield more restrictive results). None of the patients show any clear trend of increasing or decreasing classification accuracy throughout the BCI training phase. There is a substantial spread in the trial-to-trial accuracies indicating a high variability across trials within single sessions for both patients. Classification accuracy is calculated from single trial accuracy. This high variability indicates that the patients were able to successfully perform MI and achieve high classification accuracy on a portion of trials as well as failing on the other portion of trials.
